# Joint estimation and imputation of variant functional effects using high throughput assay data

**DOI:** 10.1101/2023.01.06.23284280

**Authors:** Tian Yu, James D. Fife, Ivan Adzhubey, Richard Sherwood, Christopher A. Cassa

## Abstract

Deep mutational scanning assays enable the functional assessment of variants in high throughput. Phenotypic measurements from these assays are broadly concordant with clinical outcomes but are prone to noise at the individual variant level. We develop a framework to exploit related measurements within and across experimental assays to jointly estimate variant impact. Drawing from a large corpus of deep mutational scanning data, we collectively estimate the mean functional effect per AA residue position within each gene, normalize observed functional effects by substitution type, and make estimates for individual allelic variants with a pipeline called FUSE (**Fu**nctional **S**ubstitution **E**stimation). FUSE improves the correlation of functional screening datasets covering the same variants, better separates estimated functional impacts for known pathogenic and benign variants (ClinVar *BRCA1*, p=2.24×10^−51^), and increases the number of variants for which predictions can be made (2,741 to 10,347) by inferring additional variant effects for substitutions not experimentally screened. For UK Biobank patients who carry a rare variant in *TP53*, FUSE significantly improves the separation of patients who develop cancer syndromes from those without cancer (p=1.77×10^−6^). These approaches promise to improve estimates of variant impact and broaden the utility of screening data generated from functional assays.

**Graphical Abstract:** 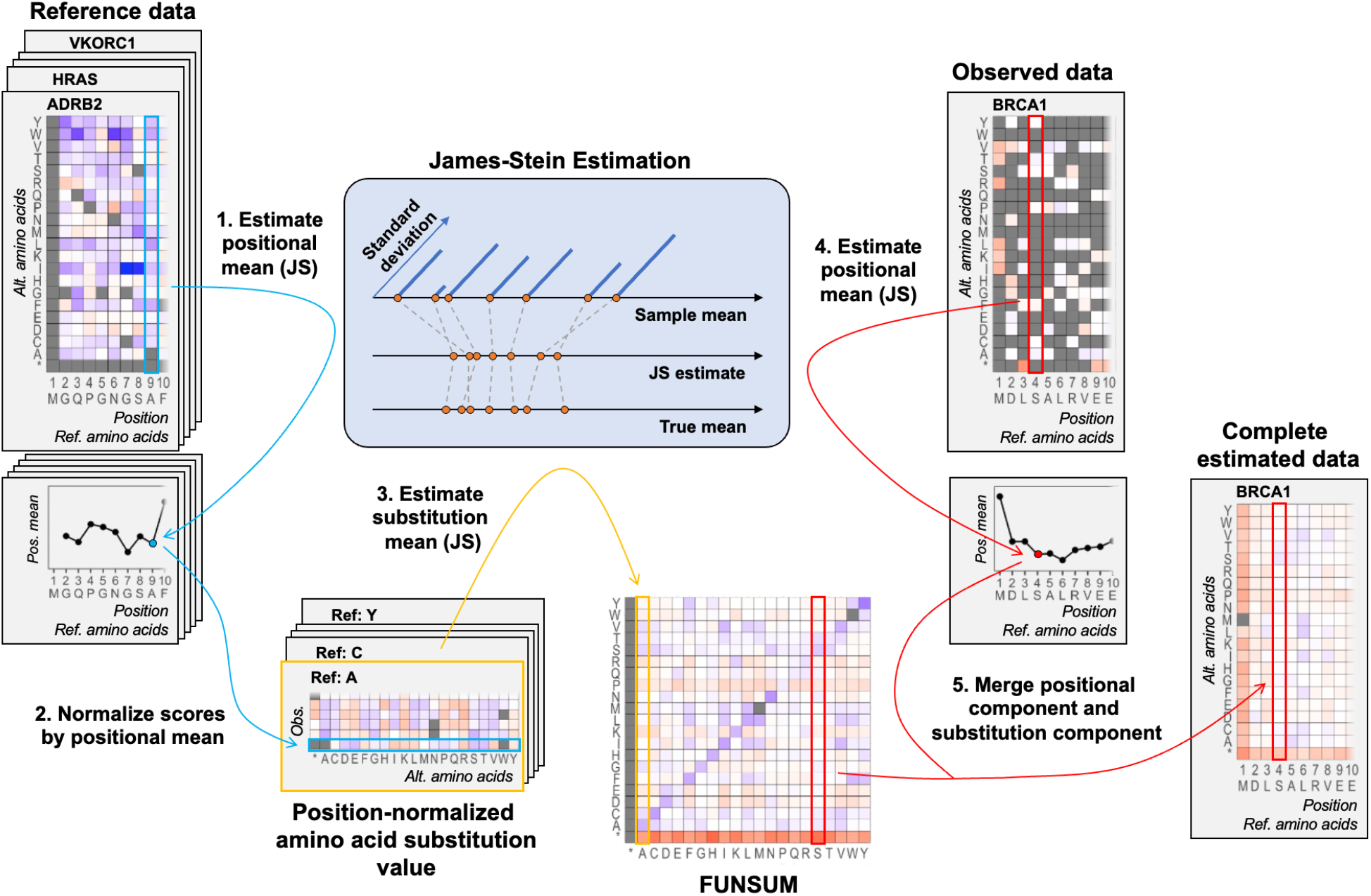

**Highlights:**

- Uses functional assay data collectively to improve the estimation of allelic variant effects
- Infers the impact of variants not experimentally screened, broadening the utility of assays
- Improves the discrimination of clinically actionable variants within ClinVar
- Significantly separates patients at risk for cancer syndromes in the UK Biobank

## Introduction

Mapping germline variants to personalized clinical risk is a major goal in precision medicine.^1^ While clinical sequencing has advanced dramatically, the interpretation of monogenic variants remains challenging, even in established disease genes.^2^ Despite extensive diagnostic testing, most variants have only been observed in a few cases or controls, if at all.^3^ As a result, the vast majority are classified as Variants of Uncertain Significance (VUS), preventing their use in clinical management,^4^ as disclosure guidelines recommend such results not be communicated to patients and providers.^5^ Extensive prior work to estimate variant functional effects includes the use of evolutionary conservation data,^6–8^ structural features,^9^ and aggregated assay data to measure the concordance and relative effects across substitution types and contexts.^10^

Deep mutational scanning (DMS) assays enable the functional assessment of thousands of coding variants which are installed in a cell line, typically replacing the native gene sequence.^11–17^ These *in vitro* assay results are highly concordant with clinical phenotype data, and such estimates have been used to systematically resolve VUSs in genes associated with clinically actionable syndromes.^18,19^ One study which used DMS data to classify variants in *BRCA1* found that functional evidence could be used to reclassify 11% of VUS as pathogenic (297/2,701).^14^ The ACMG/AMP sequence variant interpretation guidelines specify that ‘well-established’ functional studies are considered strong sources of evidence (criteria PS3/BS3).^2^ Methods to improve the utility of functional data could help resolve the large number of VUSs in variant databases such as ClinVar.^20^

The emergence of precise CRISPR-Cas9 techniques such as base and prime editing has begun to increase the scale of functional screening dramatically, allowing interrogation of variants across many genes.^15,21,22^ However, individual predictions of functional effect are prone to sources of statistical noise due to variance in assay readouts or editing efficiency.^23^ In addition, some approaches such as base editing cannot yet experimentally measure functional effects of all possible missense variants, leading to incomplete assay coverage of clinically important variants.

Here, we develop a framework to improve the estimation of variant functional effects by integrating data across experimental observations. Given that there are often many related measurements in high-throughput screens, we make use of the entirety of the data to identify sources of correlated noise in order to improve the estimation of functional impacts. We find that this approach can significantly improve the concordance of functional assays covering the same substitutions, improve our ability to discriminate between established pathogenic and benign variation, and can significantly separate patients who may be at increased clinical risk in population cohorts.

## Results

We developed FUSE (**Fu**nctional **S**ubstitution **E**stimation), a joint estimation pipeline that collectively uses assay data to improve the accuracy of functional estimates. The FUSE pipeline reduces variance inherent in experimental assays, drawing from related observations of mutational effects at each AA residue, and throughout each protein. FUSE also imputes values for the functional impact of a substantially larger set of amino acid substitutions when assay data is incomplete (**Graphical Abstract, Methods**).

FUSE draws from a library of comprehensive DMS datasets in human genes. It first estimates the mean functional effect of variation at each AA residue (‘positional mean effect’) using shrinkage estimation (James-Stein Estimator, JSE), which provides a baseline positional estimate of the expected impact for any amino acid substitution in each residue. Next, FUSE uses these mean positional scores to normalize the functional effects for each amino acid substitution (‘substitution effect’), resulting in a functional substitution matrix (FUNSUM). Finally, to make estimates of functional impact for individual allelic variants, FUSE first estimates the mean positional effect at each AA residue from any observed data, and then adjusts the positional mean using the residual value for the specific substitution type from the FUNSUM matrix (**Methods**). We applied the FUSE pipeline to improve the estimation of functional impact for coding variants in experimental assays across a set of cancer predisposition genes, and measured its utility in separating established pathogenic and benign variants, as well as rare missense variants in a large patient cohort.

### Generating a Functional Substitution Matrix

We developed a Functional Substitution Matrix (FUNSUM) to generalize information about observed functional effects associated with each amino acid substitution using previously generated DMS assay data. Using the approach outlined in the **Graphical Abstract**, we calculated the estimated functional effect of each substitution, adjusting for the positional mean effect within each AA residue position. We then used shrinkage estimation (JSE) to aggregate the effects of each substitution type, and combined them into the FUNSUM matrix (**Figure 1**). We found that across a broad set of cell lines and screening strategies used in 13 functional assays, the functional effects captured by FUNSUM are not driven by one gene or assay. To measure this, we generated 13 alternative FUNSUM matrices in which one assay was omitted, and conducted a leave-one-out correlation analysis. We found high correlation among the alternative matrices (**Figure S1**, R^2^>0.95). Finally, using hierarchical clustering, we identified the 9 most correlated genes and used these to develop a final FUNSUM matrix (**STAR Methods** and **Figure S2**).

**Figure 1:**
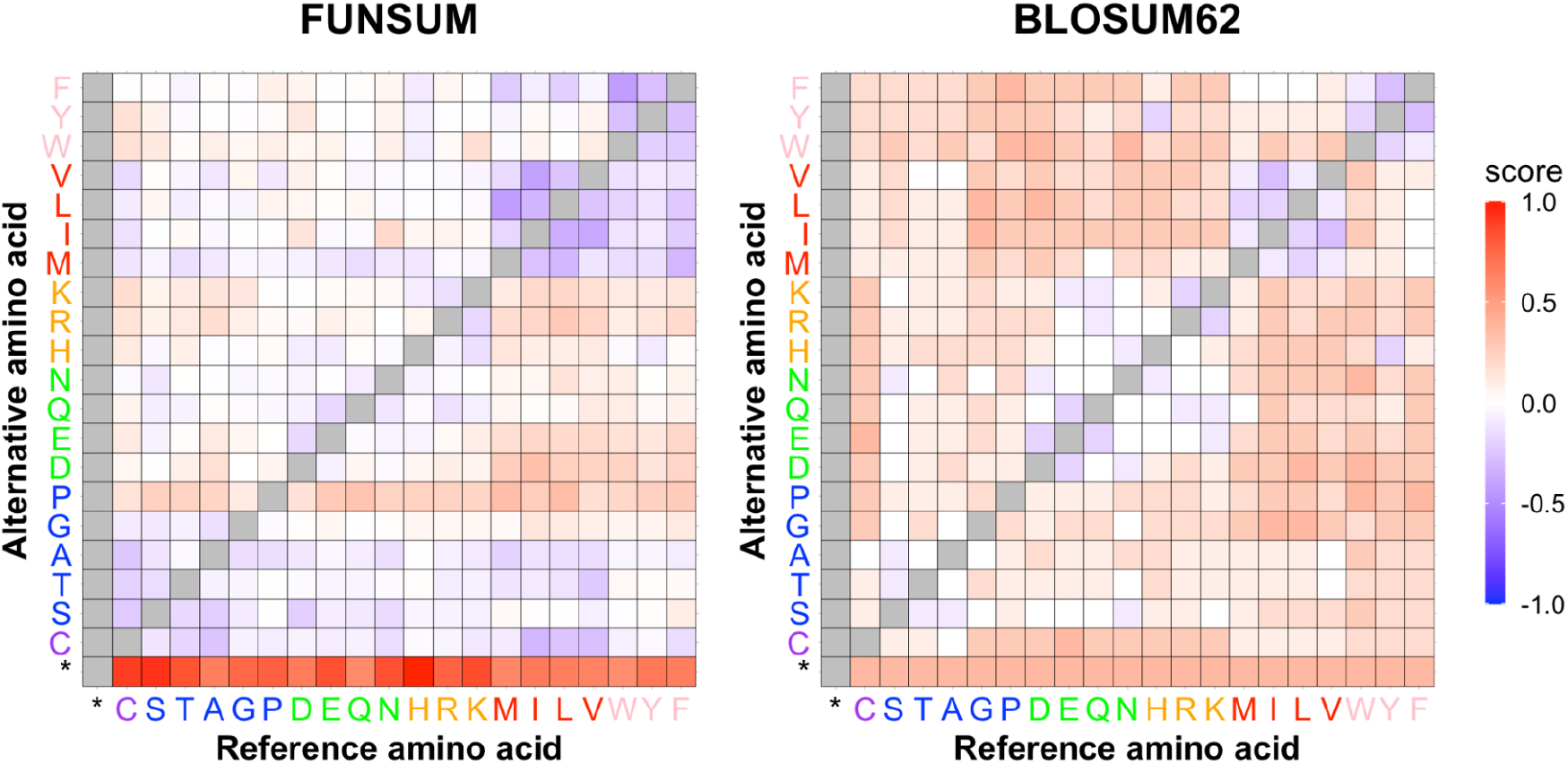
FUNSUM and BLOSUM62 substitution matrices. The FUNSUM matrix is generated by calculating the residual effects of each amino acid substitution, using a multi-stage pipeline that leverages shrinkage estimation (James-Stein estimator). For comparison purposes, both the FUNSUM and BLOSUM62 substitution matrices were normalized by standard deviation. Higher scores reflect more deleterious substitutions, with unavailable data colored in gray. The FUNSUM matrix identifies strong functional effects from known damaging changes, including substitutions to proline or termination codons.

In contrast to the widely used BLOSUM62 matrix (rescaled for comparison), the FUNSUM substitution matrix may capture distinct information useful in functional estimation analysis. FUNSUM is asymmetric, providing estimates of functional impact for substitutions in both directions (A->V vs. V->A). The correlation between FUNSUM and BLOSUM62 is substantial (R^2^ = 0.70) but incomplete, indicating that FUNSUM is likely capturing information about amino acid substitutions distinct from the evolutionary impact reflected by BLOSUM62. We found that in estimating impacts of functional effects, FUNSUM is superior to BLOSUM62 for both natively sequenced and inferred data (**Figure S3**).

### Improving the estimation of functional effects from assay data

We first evaluated whether FUSE can improve correlation between datasets which have been experimentally screened using orthogonal approaches. A broad set of variants in *BRCA1* have been screened using both base editing^16^ and saturation genome editing profiling.^14^ A total of 192 variants were screened on both platforms, and the Pearson correlation between these originally published functional scores is 0.06 (p-value = 0.395). After processing the base editing assay results with FUSE, the correlation increased substantially to 0.33 (p-value = 4.20×10^−6^). FUSE also infers additional variants which were not natively screened on the base editing assay. A total of 749 variants overlap between the full FUSE set (including inferred sites), and inclusion of these variants increased the correlation to 0.36 (p-value to 2.2×10^−16^).

### Improving the assessment of clinically actionable variants

Next, we measured whether FUSE can significantly improve the estimation of variants with clinical assertions in ClinVar. We first evaluated functional scores for variants previously classified as pathogenic (P/LP) or benign (B/LB) with a high quality evidence base, using a set of 2,741 variants covered in a *BRCA1* base editing assay.^16^ Prior to using FUSE, the original functional scores significantly separated pathogenic variants from benign (**Figure 2A**, Kolmogorov-Smirnov [KS], p=0.00235). After refining functional estimates on the same set of 2,741 variants using FUSE, we observed greater significance in separation between pathogenic and benign variant groups (**Figure 2D**, KS p=1.22×10^−26^). We were also able to expand our estimated functional scores to a total of 10,347 variants (including 7,606 not directly assayed) by estimating the positional mean at each AA residue and adjusting it to make allelic estimates using FUNSUM (**Figure 2F**). Pathogenic variants in this expanded set of estimated functional scores were significantly different from benign variants (**Figure 2E**, KS p=7.66×10^−72^).

**Figure 2:**
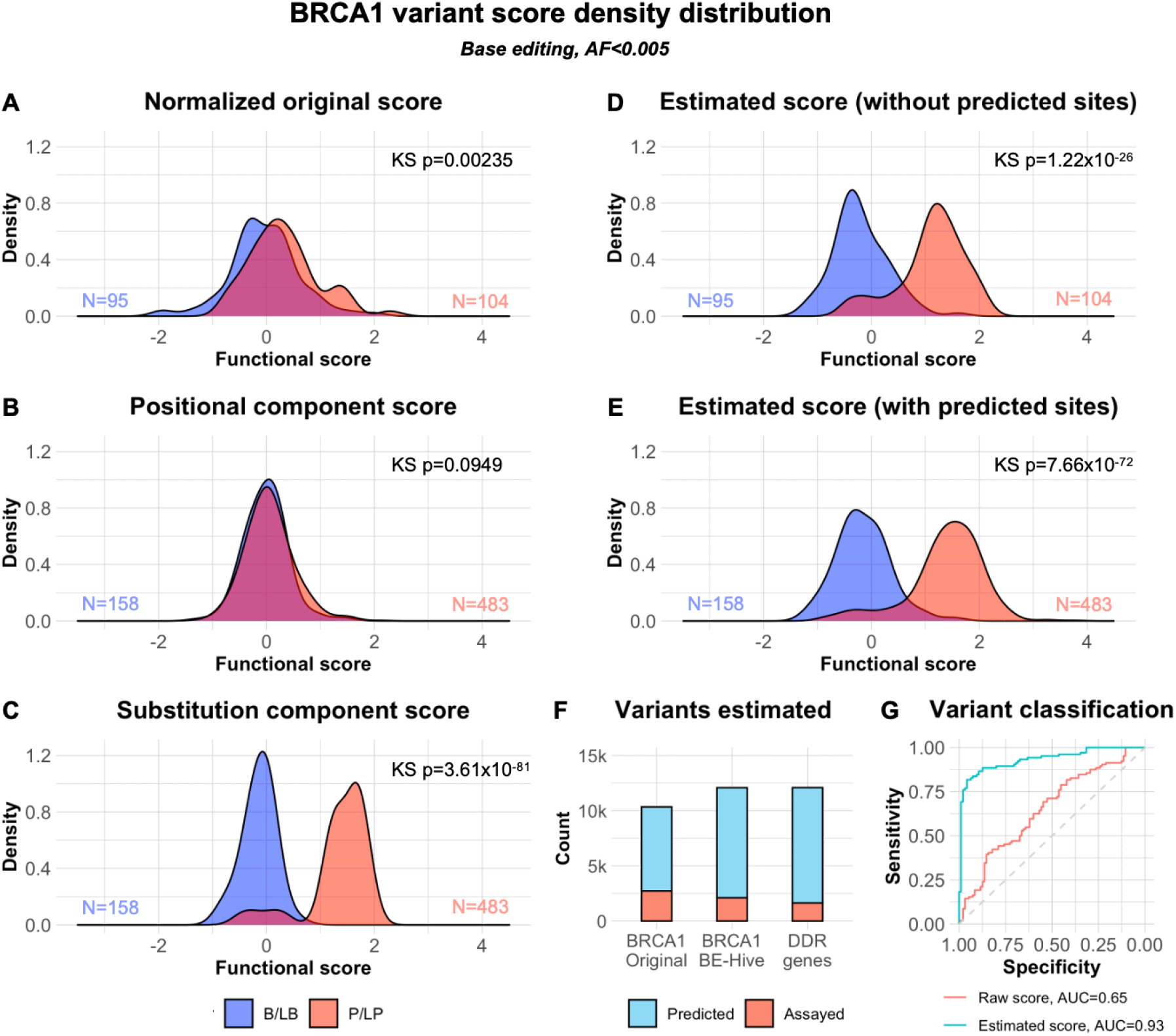
Distribution of estimated functional impact from base editing data for clinically significant variants. Clinically significant variants were categorized into benign/likely benign group (B/LB, blue) and pathogenic/likely pathogenic group (P/LP, red) based on ClinVar annotations. For each score component below, the Kolmogorov–Smirnov (KS-test) is used to measure the significance of difference between the B/LB and the P/LP groups. **[A]** Normalized variant functional scores from a published *BRCA1* base editing dataset. **[B]** Positional mean effect as estimated by JSE, the mean effect of any substitution at the AA residue position. **[C]** Substitution impact estimated from the FUNSUM amino acid substitution matrix. **[D]** FUSE estimated functional scores for variants in the original assay. **[E]** FUSE estimated functional scores for variants that were assayed or can be predicted. **[F]** The number of assayed and predicted variants with functional impact score estimated by FUSE. **[G]** Receiver operating characteristic curve (ROC) for classification of variants with existing clinical assessments (ClinVar). Logistic regression models were trained on the functional scores of pathogenic and benign variants in the DMS datasets of 10 genes, and were used to classify pathogenic and benign variants in a separate *BRCA1* base editing dataset. One model was trained and tested on functional scores from the original assay (red), and the other model was trained and tested on the functional score estimated by FUSE.

To evaluate whether FUSE could improve the classification of actionable variants, we drew from variants with existing classifications in ClinVar in *BRCA1*. First, we trained two logistic regression models using pathogenic and benign variants in 9 DMS dataset genes (not including *BRCA1*), using functional scores from the original assays and functional scores estimated by FUSE. Then, we evaluated the classification performance of these two models on a *BRCA1* base editing dataset using the original functional score and FUSE estimates, respectively. The model trained and tested on the FUSE estimated functional scores significantly outperformed the other, demonstrating the applicability of functional estimation to predict variant clinical significance (**Figure 2G**, AUC=0.93).

Base editing efficiency and the frequencies of edited outcomes can differ depending on sequence context. We used BE-Hive^23^ to predict the outcomes of edited variants and their associated probabilities for all gRNAs in the same *BRCA1* base editing assay, restricting our analysis to 2,113 edited variants with over 50% outcome probability. Using the BE-Hive pipeline and FUSE together, we estimated functional scores for 12,088 variants, including 9,975 variants which were not directly assayed. Among these variants, we observed greater separation between the P/LP and B/LB variant groups with functional scores estimated by FUSE (**Figure S4B**, KS p=3.28×10^−77^). Furthermore, we were able to improve the separation of P/LP and B/LB variants in a broader multi-gene base editing assay covering DNA Damage Response (DDR) genes (**Figure S4C**, KS p=2.9×10^−5^) from the original functional scores (KS p=0.0884), dramatically increasing the number of variants with functional scores from the original 1,643 variants to 12,096 variants.

To measure the utility of the FUSE pipeline on different functional screening approaches, we estimated effects in a DMS assay covering variants in *TP53*^17^ and a saturation genome editing assay of *BRCA1*.^14^ In the *TP53* assay, we observed slightly increased score separation at the margins for some variants, but found similar overall significance of separation using our estimated functional scores (**Figure 3D**, KS p=2.44×10^−45^) when compared with the original scores (**Figure 3A** KS p=1.38×10^−43^). In the *BRCA1* assay, we found similar results with FUSE separating variant groups (**Figure S5B**, KS p=3.55×10^−20^) compared with the original scores (KS p=4.11×10^−22^), noting that there are very few B/LB variants (n=31) overlapping with the functional dataset. Finally, we estimated functional impacts using DMS data across 10 genes, and again found similar significance in separating the P/LP and B/LB variant groups (**Figure S5C**, KS, p=8.41×10^−58^, training described in **STAR Methods**) compared to the original functional scores (KS, p=7.47×10^−56^). These assays have nearly complete coverage of possible coding variants, so they had a limited number of additional sites imputed by the FUSE pipeline.

**Figure 3:**
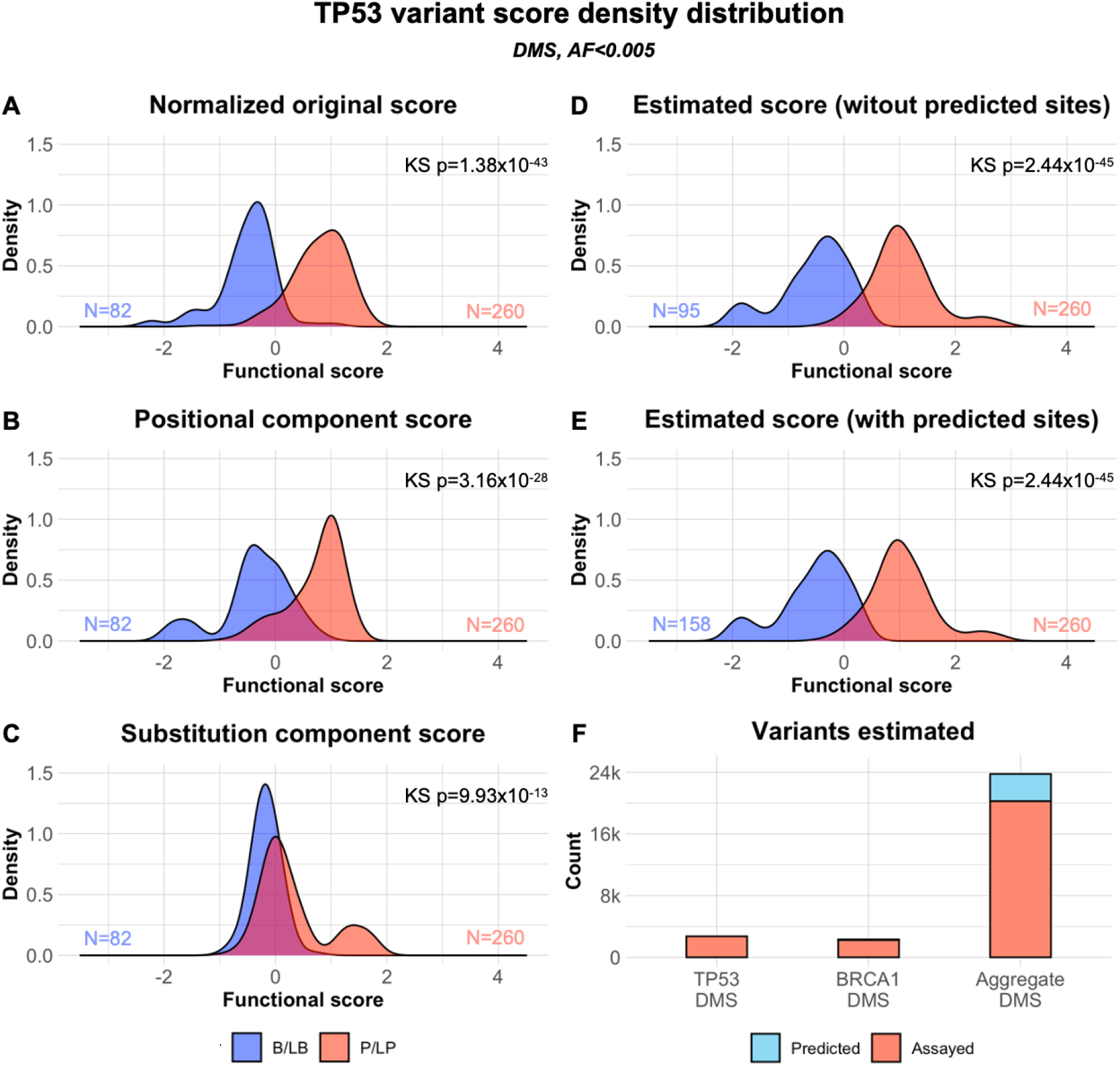
Density distribution of estimated functional impact from deep mutational scanning (DMS) data for clinically significant variants. Clinically significant variants were categorized into benign/likely benign group (B/LB, blue) and pathogenic/likely pathogenic group (P/LP, red) based on ClinVar annotations. For each score component below, the Kolmogorov–Smirnov (KS-test) is used to measure the significance of difference between the B/LB and the P/LP groups. **[A]** Normalized variant functional scores from a published *TP53* DMS dataset. **[B]** Positional mean effect as estimated by JSE, the mean effect of any substitution at the AA residue position. **[C]** Substitution impact estimated from the FUNSUM amino acid substitution matrix. **[D]** FUSE estimated functional scores for variants in the original assay. **[E]** FUSE estimated functional scores for variants that were assayed or can be predicted. **[F]** The number of assayed and predicted variants with functional impact score estimated by FUSE.

### Validation using patient cancer outcomes in the UK Biobank

Next, we evaluated whether the FUSE pipeline can separate carriers of rare missense variants at increased clinical risk of cancer in the UK Biobank.^24^ We first used the FUSE pipeline on functional data from *TP53* and *BRCA1* assays, blinded to patient phenotypic status (**Methods**), and then assessed the significance of separation of patients who develop or do not develop related cancer syndromes. In a *TP53* DMS assay, functional impacts estimated by FUSE were better at separating patients who develop Li-Fraumeni syndrome (LFS) related cancer syndromes from those without LFS-related cancer syndromes (**Figure 4D**, p=1.77×10^−6^) when compared with the original functional scores (**Figure 4A**, p=2.26×10^−4^). In a *BRCA1* saturation genome editing assay, the pipeline better separated patients who developed early-onset breast cancer from those without breast cancer (**Figure S6A**, KS p=0.025). Similarly, in a *BRCA1* base editing assay, we found that FUSE increased the number of carriers for whom predictions can be made from functional data (803 to 2,680), and improved the separation between early breast cancer patients from non-breast cancer patients (**Figure S6D**, KS p=0.00993).

**Figure 4:**
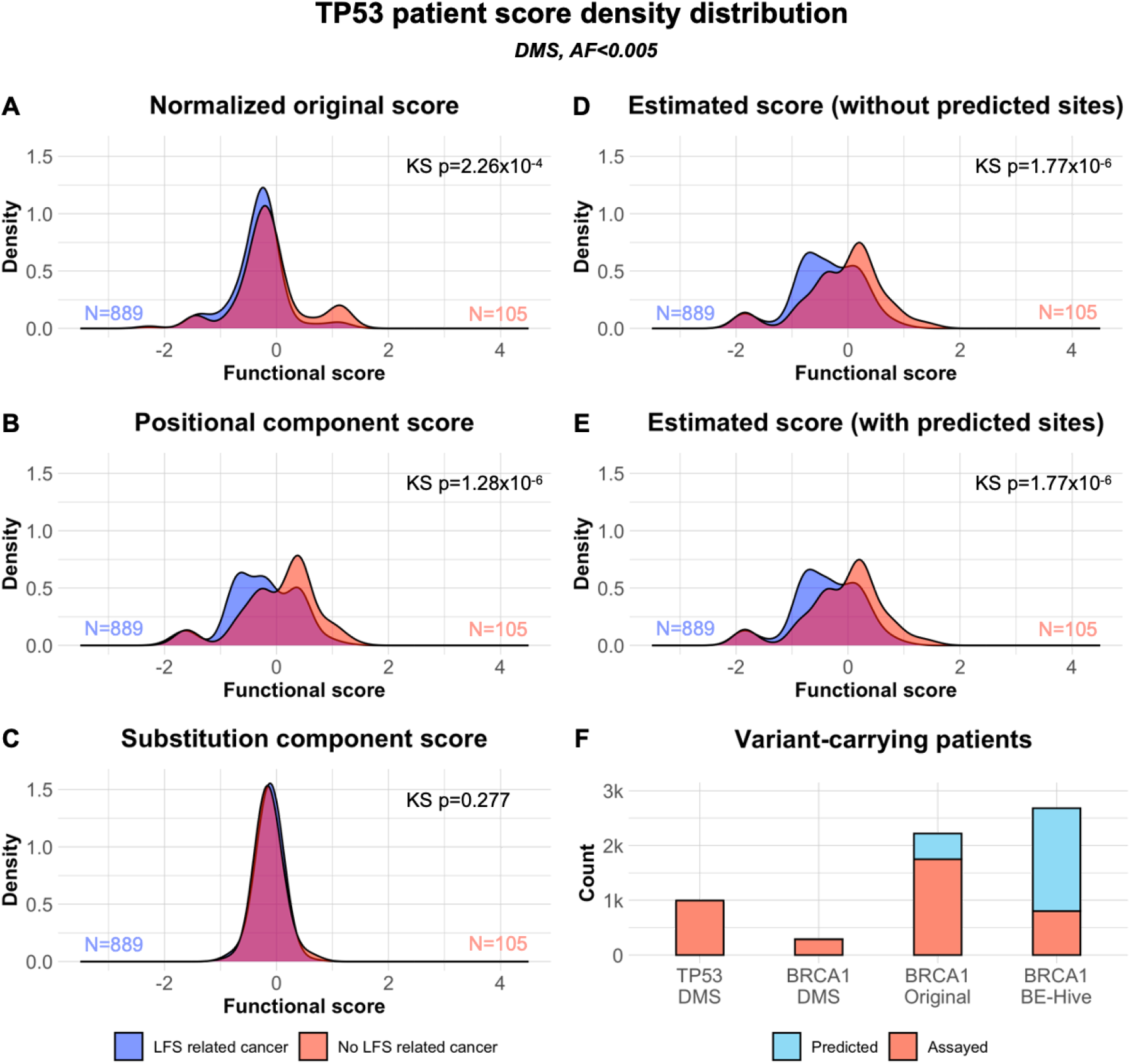
Density distribution of estimated functional impact of variants in patient groups categorized by UKBB patient characteristics. The patients who carried TP53 variants were categorized by whether they had Li-Fraumeni syndrome associated cancers (LFS, red), or did not have Li-Fraumeni syndrome associated cancers (Non-LFS, blue). The patient score was represented by the functional score of the TP53 variants detected. Patients with multiple TP53 variants were very rare and not included. The numbers of patients in each group are indicated by their corresponding color. For each score component below, the Kolmogorov–Smirnov (KS-test) is used to measure the significance of difference between the non-cancer and cancer groups. **[A]** Normalized variant functional scores from a published *TP53* DMS dataset. **[B]** Positional mean effect as estimated by JSE. **[C]** Substitution impact estimated from the FUNSUM amino acid substitution matrix. **[D]** FUSE estimated functional scores for variants in the original assay. **[E]** FUSE estimated functional scores for variants that were assayed or can be predicted. **[F]** The number of patients that carry variants with functional impact score assayed or predicted by FUSE.

## Discussion

Experimental assays provide a unique lens into the functional effects of missense variants. Prior methods have used deep mutational scanning data to resolve VUSs, to estimate the functional effects of amino acid substitution types, and to inform library design.^23,28^ While functional assays are excellent proxies of the impact of variants, these measurements may be individually noisy. As deep mutational scanning methods generate nearly complete information about substitutions in a protein region, there is a great deal of correlated information. FUSE takes advantage of such repeated and related measurements to reduce variance and improve estimates of functional effect. Additionally, screening techniques such as base editing assays can only install a subset of possible coding variants. FUSE is able to make estimates for these variants which could not be experimentally screened, dramatically expanding the overlap with clinical variant datasets.

In the clinical domain, reclassifying VUSs is a critical translational gap: for most patients who have no clinical indication for testing (those prospectively screened, or in biobanks), current practice guidelines discourage the reporting of VUSs in the context of screening or secondary findings,^5^ even though they may confer increased risk. However, it is still challenging to determine which are likely to be highly impactful versus those which may have limited or no effects. When patients and clinicians have incomplete or incorrect information, it can result in serious clinical and/or sociological consequences, including unnecessary surveillance or irreversible prophylactic surgery.^25,26^ In one retrospective study of *BRCA1* or *BRCA2* VUS carriers, 39% of individuals without cancer opted to receive a bilateral mastectomy, and among all patients with and without cancer, and 21 of 97 or 22% of patients had a VUS reclassified, with 95% downgraded to benign.^27^ While the evidence base has significantly improved with extensive diagnostic sequencing and expert review panels, resolving the many outstanding VUS classifications remains an important goal.

At present, there are a limited number of comprehensive DMS datasets for human genes. These datasets were ascertained due to their clinical importance for a set of phenotypes, which could affect the generalizability of these results. As additional screening data become available in public repositories such as MaveDB and through screening consortia such as IGVF (Impact of Genomic Variation on Function), we expect the quality and generalizability of FUNSUM and FUSE to improve.

There are also challenges in integrating multiple datasets into a single functional substitution matrix. Given that functional screens use different experimental platforms, screening strategies, phenotypic readouts, and experimental conditions (e.g., treatment groups), the ability to combine these datasets may pose challenges. Furthermore, there may be differing functional effects across proteins or protein domains and regions, which may require the pre-selection of regions which are likely to have functional impact, such as regions under negative selection, experimentally identified as deleterious, or statistically enriched in cases. Finally, though the FUSE pipeline improves the global estimation of variants (JSE has been shown to dominate the maximum likelihood estimator (MLE) in terms of total squared error), JSE is a biased estimator, which could negatively affect individual estimates which are indeed outliers. The larger the variance is within any population of variants, the more biased the estimation of the population mean will be towards the global mean.

Future work may focus on the integration of computational predictions of variant effect with FUSE estimates, and in rational library design which makes use of this pipeline, as the search space of all possible non-synonymous variants is intractably large for even the most high-throughput approaches. We demonstrated that the FUSE estimation pipeline improves correlation among functional studies, better separates variants which have been previously classified as pathogenic or benign, and significantly separates variation in individuals with and without disorders. In conclusion, this approach promises to improve the quality and broaden the utility of data generated from deep mutational scanning assays.

## STAR★Methods

### Key resources table

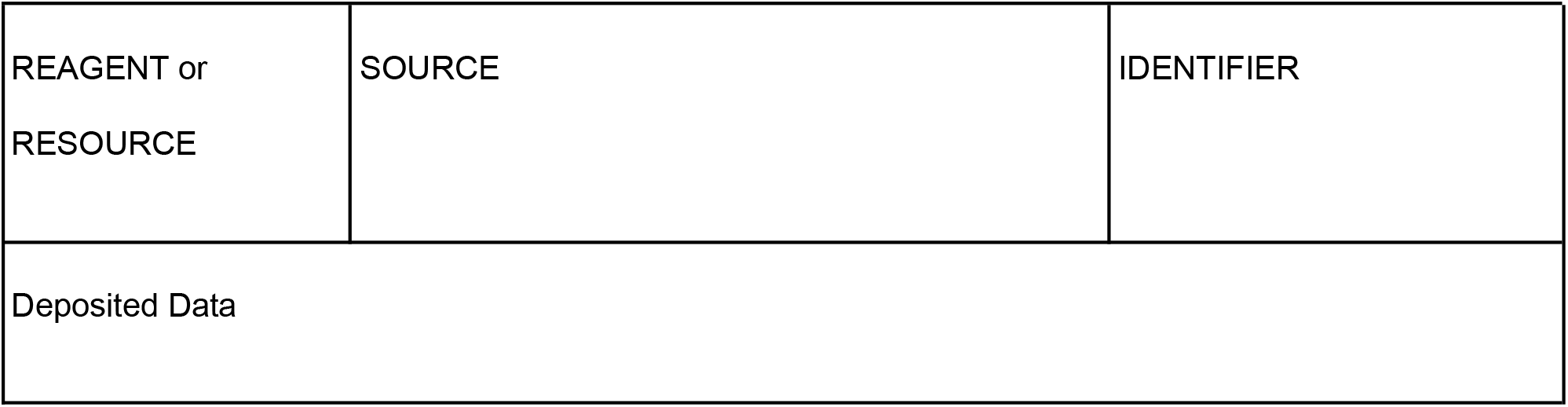
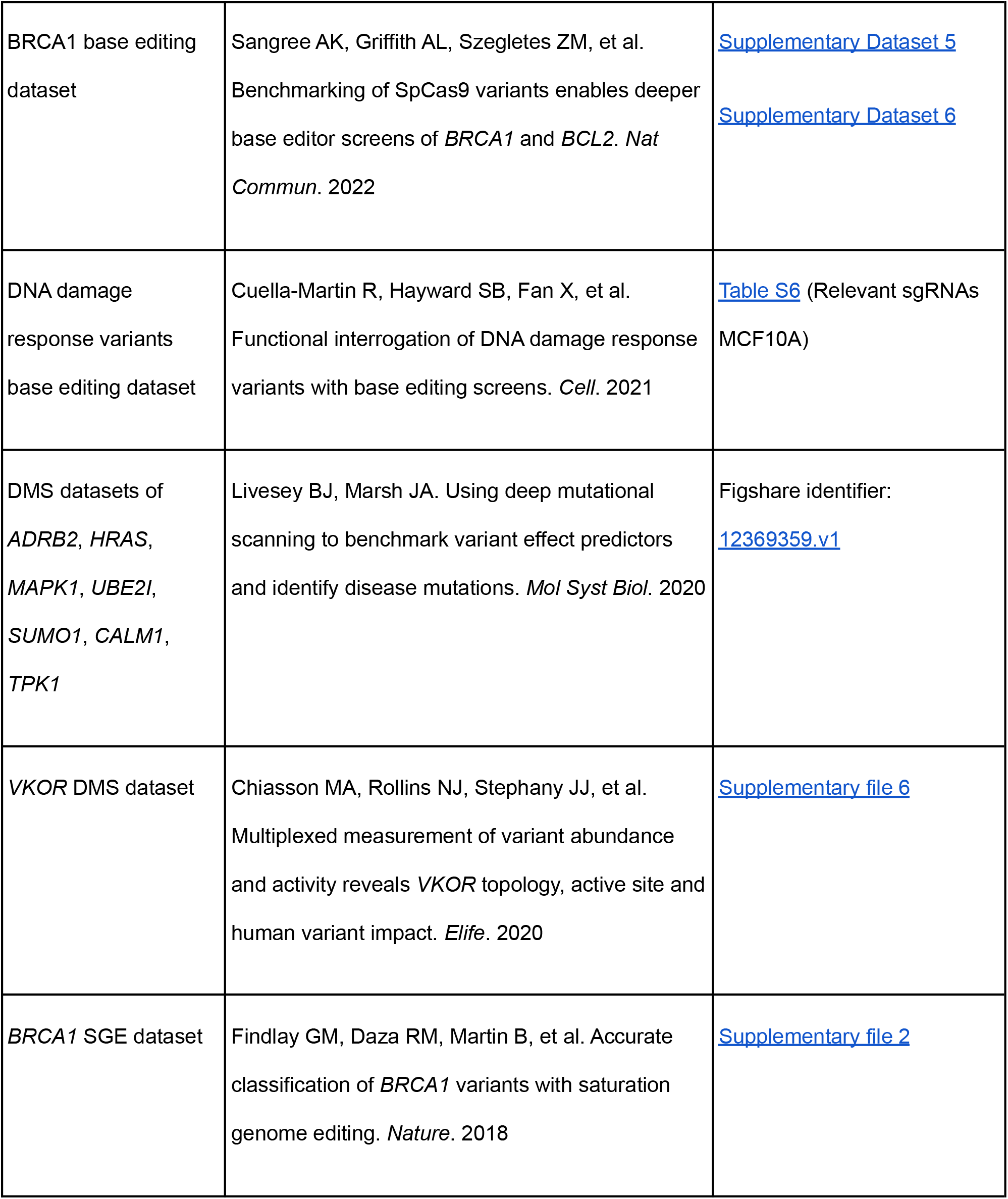
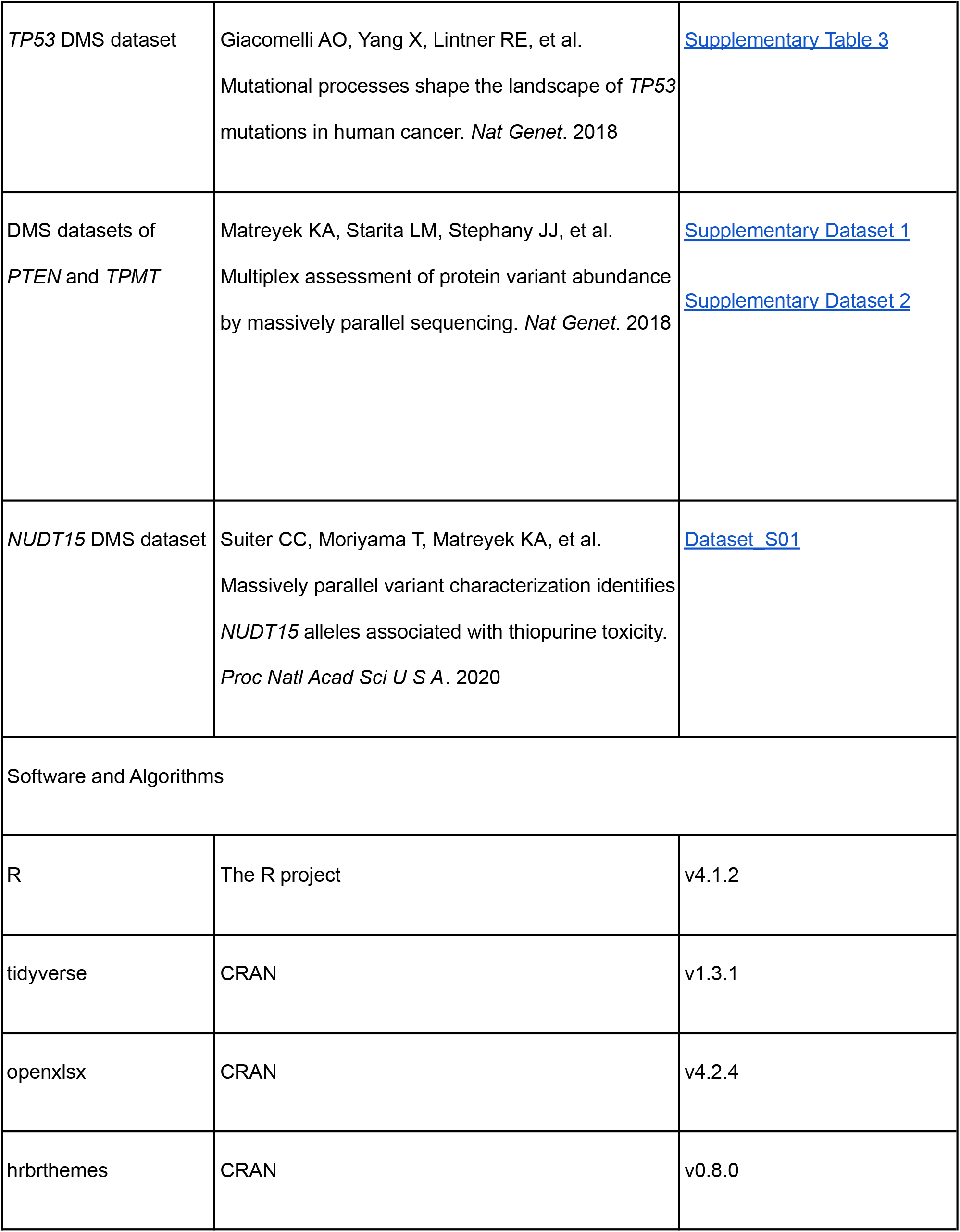
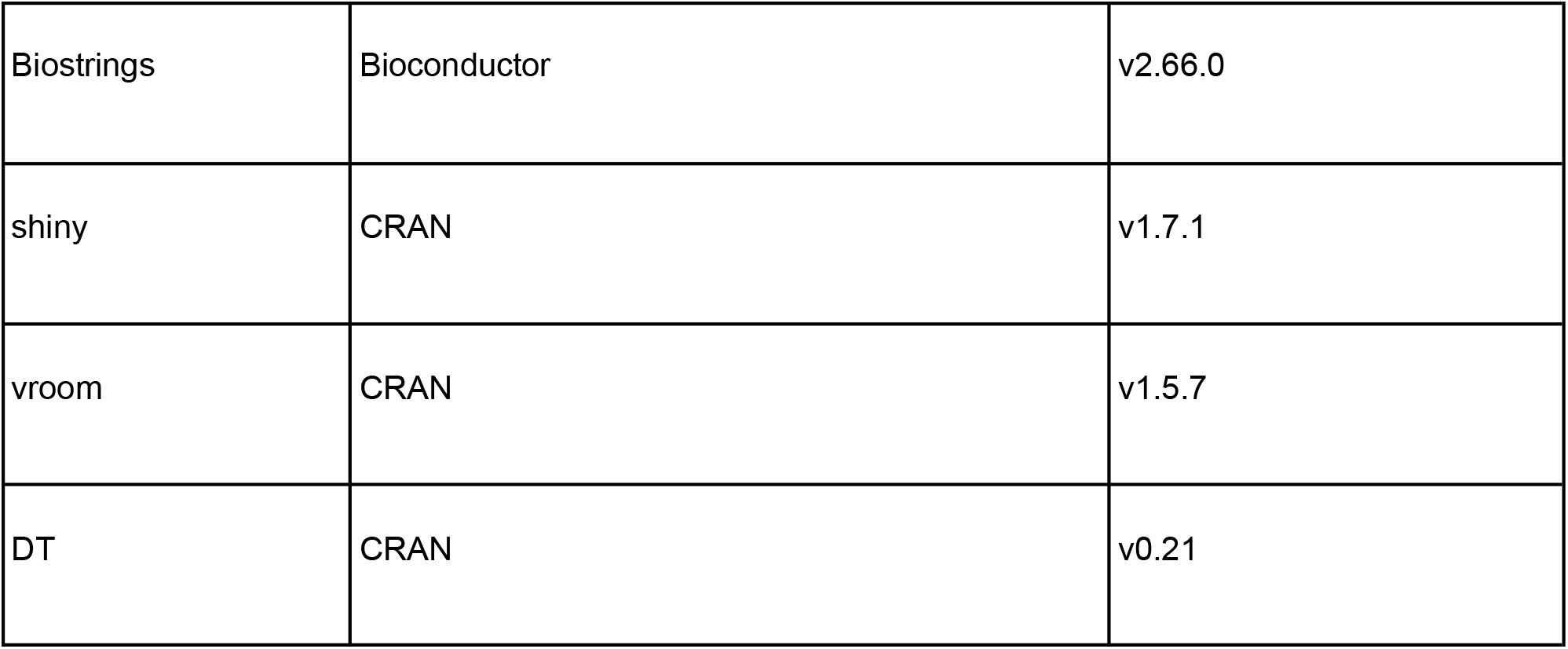

| REAGENT or RESOURCE | SOURCE | IDENTIFIER |
| --- | --- | --- |
| Deposited Data |  |  |
| BRCA1 base editing dataset | Sangree AK, Griffith AL, Szegletes ZM, et al.<br>Benchmarking of SpCas9 variants enables deeper base editor screens of *BRCA1* and *BCL2*. *Nat Commun.* 2022 | Supplementary Dataset 5<br>Supplementary Dataset 6 |
| DNA damage response variants base editing dataset | Cuella-Martin R, Hayward SB, Fan X, et al.<br>Functional interrogation of DNA damage response variants with base editing screens. *Cell.* 2021 | Table S6 (Relevant sgRNAs MCF10A) |
| DMS datasets of *ADRB2*, *HRAS*, *MAPK1*, *UBE2I*, *SUMO1*, *CALM1*, *TPK1* | Livesey BJ, Marsh JA. Using deep mutational scanning to benchmark variant effect predictors and identify disease mutations. *Mol Syst Biol.* 2020 | Figshare identifier: 12369359.v1 |
| VKOR DMS dataset | Chiasson MA, Rollins NJ, Stephany JJ, et al.<br>Multiplexed measurement of variant abundance and activity reveals *VKOR* topology, active site and human variant impact. *Elife.* 2020 | Supplementary file 6 |
| *BRCA1* SGE dataset | Findlay GM, Daza RM, Martin B, et al. Accurate classification of *BRCA1* variants with saturation genome editing. *Nature.* 2018 | Supplementary file 2 |
| TP53 DMS dataset | Giacomelli AO, Yang X, Lintner RE, et al.<br>Mutational processes shape the landscape of *TP53* mutations in human cancer. *Nat Genet.* 2018 | Supplementary Table 3 |
| DMS datasets of *PTEN* and *TPMT* | Matreyek KA, Starita LM, Stephany JJ, et al.<br>Multiplex assessment of protein variant abundance by massively parallel sequencing. *Nat Genet.* 2018 | Supplementary Dataset 1<br>Supplementary Dataset 2 |
| *NUDT15* DMS dataset | Suiter CC, Moriyama T, Matreyek KA, et al.<br>Massively parallel variant characterization identifies *NUDT15* alleles associated with thiopurine toxicity. *Proc Natl Acad Sci U S A.* 2020 | Dataset_S01 |
| Software and Algorithms |  |  |
| R | The R project | v4.1.2 |
| tidyverse | CRAN | v1.3.1 |
| openxlsx | CRAN | v4.2.4 |
| hrbrthemes | CRAN | v0.8.0 |
| Biostrings | Bioconductor | v2.66.0 |
| shiny | CRAN | v1.7.1 |
| vroom | CRAN | v1.5.7 |
| DT | CRAN | v0.21 |

### Resource availability

#### Lead contact

Further information and requests should be directed to and will be fulfilled by the lead contact, Christopher Cassa.

#### Materials availability

This study did not generate new unique reagents.

#### Experimental model and subject details

No experimental models were utilized as part of this publication. No novel subjects were collected as part of this publication.

#### Method details

##### Selection and pre-processing of existing deep mutational scanning datasets

We make use of prior deep mutational scanning (DMS) assays generated across a variety of genes which span a variety of experimental platforms and cell types^11–13,29^ as input for building a functional substitution matrix (FUNSUM), on the basis of their nearly-complete coverage of possible amino acid substitutions, and a correlation analysis of functional effects across genes (**Figure S5**). DMS and base editing data from *BRCA1*, and DMS data from *TP53* were held aside for validation purposes, given their clinical significance and the many variants in these genes annotated in ClinVar.

To make use of the data collectively, we normalize functional effects across experiments. For each DMS dataset, median standardization was used (functional scores were standardized by subtracting the median functional score and then dividing by the standard deviation of all scores.) Next, we use known pathogenic and benign variants from ClinVar to align the direction of effect of the functional score to ensure the score of mean pathogenic variants is greater than the mean score of benign variants. In genes where benign variants were not present in ClinVar, the mean of all variant scores was used in place to align the direction of DMS scores.

##### FUSE pipeline overview

The functional effect of an amino acid substitution can be influenced by both the position of the amino acid within the protein, as well as the impact of the specific substitution. Thus, we dissect the functional effect into a positional component and a substitution component. The positional component can be estimated as the mean functional effect of all possible coding non-synonymous substitutions at a given AA residue position, when accounting for the specific substitutions which are measured. The substitution component represents the remaining expected variation in functional effect after being positionally normalized.

The estimation pipeline makes use of the James-Stein estimator (JSE), a shrinkage estimator that brings the sample mean of individual populations closer to the global mean of all populations. The pipeline has several stages: 1) using a large set of previously generated deep mutational scanning (DMS) data, we estimate the positional mean value for mutations at each AA residue in 8 genes, which is then used to 2) calculate the positionally-normalized activity of each possible observed amino acid substitution across the genes, 3) using these normalized observations, we then estimate the mean substitution value and assemble it as the functional substitution matrix (FUNSUM), which captures the impact of substitutions, regardless of the effect of gene and amino acid positions. 4) To apply this pipeline on new functional assay data, the scores need to be first normalized by median and standard deviation, and their positional means estimated by JSE, and 5) then individual scores are calculated as the sum of the positional mean estimate and the substitution scores in FUNSUM.

##### Estimating positional mean functional values from reference datasets

For each of the pre-processed DMS datasets, functional scores can be arranged into a *n* by *m* matrix, where *n* is the total number of amino acid positions with functional scores, and *m* is the 21 possible amino acid substitutions (including the stop codon) that can occur (**Figure 1**, step 1). The mean functional values for the *n* amino acid positions can be calculated simultaneously using the James-Stein Estimator (JSE). JSE considers all observed values simultaneously and outperforms MLE mean estimates in terms of total squared errors when three or more dimensional data is available.^30,31^ As n >> 3, JSE should be able to yield better positional estimates than MLE. Let *mbar* be the global mean functional score, *mu*_*MLE*_ be a vector of positional mean functional scores calculated via MLE, and *s*^*2*^ be the global variance, then the vector of positional mean functional scores estimated via JSE, *mu*_*JSE*_, can be calculated as 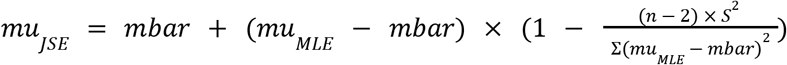.

##### Developing an amino acid substitution matrix derived from functional data

For a given amino acid position, individual functional scores can be positionally normalized by subtracting the positional mean functional value estimated via JSE (**Figure 1**, step 2). Here we aggregate the positionally normalized functional scores from all 9 reference DMS datasets, and re-organize them based on the reference (wild-type) amino acid. Functional scores associated with the same reference amino acid can be further arranged into an *n* by *m* matrix, where *n* is the 21 possible amino acid substitutions (including the stop codon) that can occur, and *m* is the total number of occurrences of the substitutions. Similarly, we use JSE to estimate the mean functional effects for the *n* amino acid substitutions simultaneously (**Figure 1**, step 3). Finally, the mean functional effects of substitutions from all possible reference amino acids can be combined to form the FUNSUM (**Fun**ctional **Su**bstitution **M**atrix) amino acid substitution matrix.

##### Making estimates of functional effect for individual allelic variants

To improve estimates for individual allelic variants in a new functional dataset, we order the functional scores into an *n* by *m* matrix, where *n* is the total number of amino acid positions with functional scores, and *m* is the 21 possible amino acid substitutions (including the stop codon) that can occur. The positional mean effect for each amino acid position is estimated using JSE (**Figure 1**, step 4). Next, for an allelic variant, we start with the positional mean value, and then add or subtract the residual value from the FUNSUM matrix, using the value for the specific amino acid substitution caused by the allelic variant. Using this approach, it is possible to infer the functional effects of variants for which we do not have primary measurements.

##### Validation using clinical diagnostic (ClinVar) and patient data (UK Biobank)

We used variants with known clinical significance in Clinvar to assess how well the functional scores of pathogenic variants can be separated from benign variants. We retrieved all known pathogenic (P/LP) and benign (B/LB) variant annotations for all genes. The distribution of normalized functional scores, the positional component scores, the substitution component scores, the improved scores, and the improved scores with predictions were compared between benign variants and pathogenic variants. The Kolmogorov-Smirnov (KS) test was used to test for significant differences between the scores of the benign group and the pathogenic group.

Patient data from UK Biobank was used to identify cancer and non-cancer patient groups related to each functional dataset, and similarly the KS test was used to quantify the distance between the variant scores of the cancer and the non-cancer groups. For both the *BRCA1* DMS dataset, we identified *BRCA1* variants that were carried by breast cancer patients and *BRCA1* variants that were carried by non-breast cancer patients. For the *BRCA1* base editing dataset, the cancer group was further filtered to include only patients less than 50 years old, and the non-cancer group only included patients more than 65 years old to test for early onset of breast cancer. For the *TP53* DMS dataset, we identified variants that were carried by patients with cancers associated with Li-Fraumeni Syndrome (LFS) and *TP53* variants that were carried by patients without LFS-associated cancers. UK Biobank recorded cancer types that were associated with LFS included skin cancer, brain cancer, adrenal cancer, non-hodgkin’s lymphoma, bone metastases, leukemia, acute myeloid leukemia, breast cancer, and fibrosarcoma.

##### Clinical diagnostic variants

We analyze all nsSNVs in ClinVar,^32^ and label variants as pathogenic, or P/LP if the Clinical Significance label is ‘Pathogenic’, ‘Likely pathogenic’, and ‘Pathogenic/Likely pathogenic’, and benign, or B/LB if the Clinical Significance label is ‘Benign’, ‘Likely benign’, and ‘Benign/Likely benign’.

##### UK Biobank: study design, setting, and participants

The UK Biobank (UKB) is a prospective cohort of over 500,000 individuals recruited between 2006 and 2010 of ages 40-69 years.^24^ 200,625 participants with exome sequencing data were included in this analysis. Analysis of the UKB data was approved by the Mass General Brigham Institutional Review Board (Protocol 2020P002093). Analysis was performed under UKB application #41250.

##### Exome sequencing and variant annotation

Exome sequencing was performed for UKB participants as previously described.^24^ Variant allele frequencies were estimated from the Genome Aggregation Database (gnomAD v2.1 exomes N=125,748). Variants were included with population maximum allele frequencies of <=0.005 (Ensembl gnomAD plugin)^33^ or if not present in gnomAD. Functional annotations are derived from Variant Effect Predictor (v106).^34^ Variant functional consequences are calculated for the canonical transcript, and include synonymous, missense, or predicted LOF variants. The predicted LOF category includes nonsense variants.

##### Clinical endpoints

The primary clinical endpoints were specific to each condition: breast cancer (BC) for *BRCA1* variants and Li-Fraumeni syndrome (LFS) for *TP53* variants. Case definitions for BC and LFS use phenocodes for each related syndrome in the UKB based on hospitalization records, cancer, and death registries, previously described at the disorder level.^35^

##### Correlation analysis for gene-specific and leave-one-out FUNSUM matrices

To measure the correlation between each gene and its associated DMS assay data, we built gene-specific FUNSUM matrices using only normalized DMS data from each gene, and measured the Pearson correlation between matrices from every possible gene pair (**Figure S2**). We found that FUNSUM matrices built using the *SUMO1, CALM1* and *TPK1* datasets had a lower correlation with the FUNSUM matrices built from DMS datasets of other genes. Accordingly, we excluded *SUMO1, CALM1*, and *TPK1* when building the final version of FUNSUM.

To confirm the functional effects which are captured by FUNSUM are not driven by any specific dataset, we conducted a leave-one-out correlation analysis using 13 alternative FUNSUM matrices, each using the DMS functional scores from 12 out of 13 genes as input. High correlation was consistently observed among the alternative FUNSUM versions and the final version of the FUNSUM (**Figure S1**, R^2^>0.95), indicating that the functional effects captured by FUNSUM are stable and not biased by any single functional assay.

##### Robustness of the pipeline to improve the estimation of variants

For each DMS dataset from the 10 genes, we estimated the functional scores using an alternative FUNSUM generated without that particular dataset. The estimated functional scores were aggregated across 10 genes, and were similarly validated on the ClinVar variants with clinical assertions. For the variants originally included in the assays, the estimated functional scores showed greater significance in separating the P/LP and B/LB variant groups (**Figure S5C**, KS, p=8.41×10^−58^) compared to the original functional scores (KS, p=7.47×10^−56^). It shows that the pipeline can consistently enhance the functional effect from the assays despite slight changes in FUNSUM.

##### Statistical Tests

We use the one-sided Kolmogorov-Smirnov test when determining differences in functional scores for each variant or patient distribution, performed using R version v4.1.2, tidyverse v1.3.1, shiny v1.7.1, vroom v1.5.7, and DT v0.21. Figures were made using R version v4.1.2, and tidyverse v1.3.1.

## Data Availability

All data produced are available online at:
figshare.com/articles/dataset/FUSE_estimated_score_figshare_xlsx/21644711

https://static-content.springer.com/esm/art%3A10.1038%2Fs41467-022-28884-7/MediaObjects/41467_2022_28884_MOESM9_ESM.xlsx

https://static-content.springer.com/esm/art%3A10.1038%2Fs41467-022-28884-7/MediaObjects/41467_2022_28884_MOESM10_ESM.xlsx

https://www.cell.com/cms/10.1016/j.cell.2021.01.041/attachment/6d9a15e7-fa20-4305-8054-99735cb3cd15/mmc6.xlsx

https://figshare.com/articles/dataset/Raw_variant_effect_predictions_and_DMS_data_for_benchmarking_variant_effect_predictors_/12369359/1

https://elifesciences.org/download/aHR0cHM6Ly9jZG4uZWxpZmVzY2llbmNlcy5vcmcvYXJ0aWNsZXMvNTgwMjYvZWxpZmUtNTgwMjYtc3VwcDYtdjEuY3N2/elife-58026-supp6-v1.csv?_hash=K9Cej66q0D8H2rZuLN3Hm6VpMVjMsa7clkQm015qZEI%3D

https://www.ncbi.nlm.nih.gov/pmc/articles/PMC6181777/bin/NIHMS1501643-supplement-2.xlsx

https://www.ncbi.nlm.nih.gov/pmc/articles/PMC6168352/bin/NIHMS1501645-supplement-5.xlsx

https://static-content.springer.com/esm/art%3A10.1038%2Fs41588-018-0122-z/MediaObjects/41588_2018_122_MOESM3_ESM.txt

https://static-content.springer.com/esm/art%3A10.1038%2Fs41588-018-0122-z/MediaObjects/41588_2018_122_MOESM4_ESM.txt

https://www.pnas.org/doi/suppl/10.1073/pnas.1915680117/suppl_file/pnas.1915680117.sd01.xlsx

## Data and code availability

Pretrained models, usage examples, and documentation of downloadable code can be found on GitHub at github.com/cassalab/fuse. The online version of FUSE is available at https://tyu7.shinyapps.io/FUSE. Fully downloadable processed scores for each variant used in this study is available at figshare.com/articles/dataset/FUSE_estimated_score_figshare_xlsx/21644711.

## Acknowledgements

We gratefully acknowledge support from the National Human Genome Research Institute (R01HG010372; TY, IA, JF, CC). We are indebted to the UK Biobank and its participants who provided biological samples and data for this analysis. Work was performed under UK Biobank application #41250 and Mass General Brigham IRB protocol 2020P002093. We are also grateful for advice from Dr. Shamil Sunyaev (Harvard Medical School), Dr. Natasha Strande (Geisinger Health System), Vineel Bhat, and Nana Twumasi-Ankrah.

## Supplementary Figures

**Figure S1:**
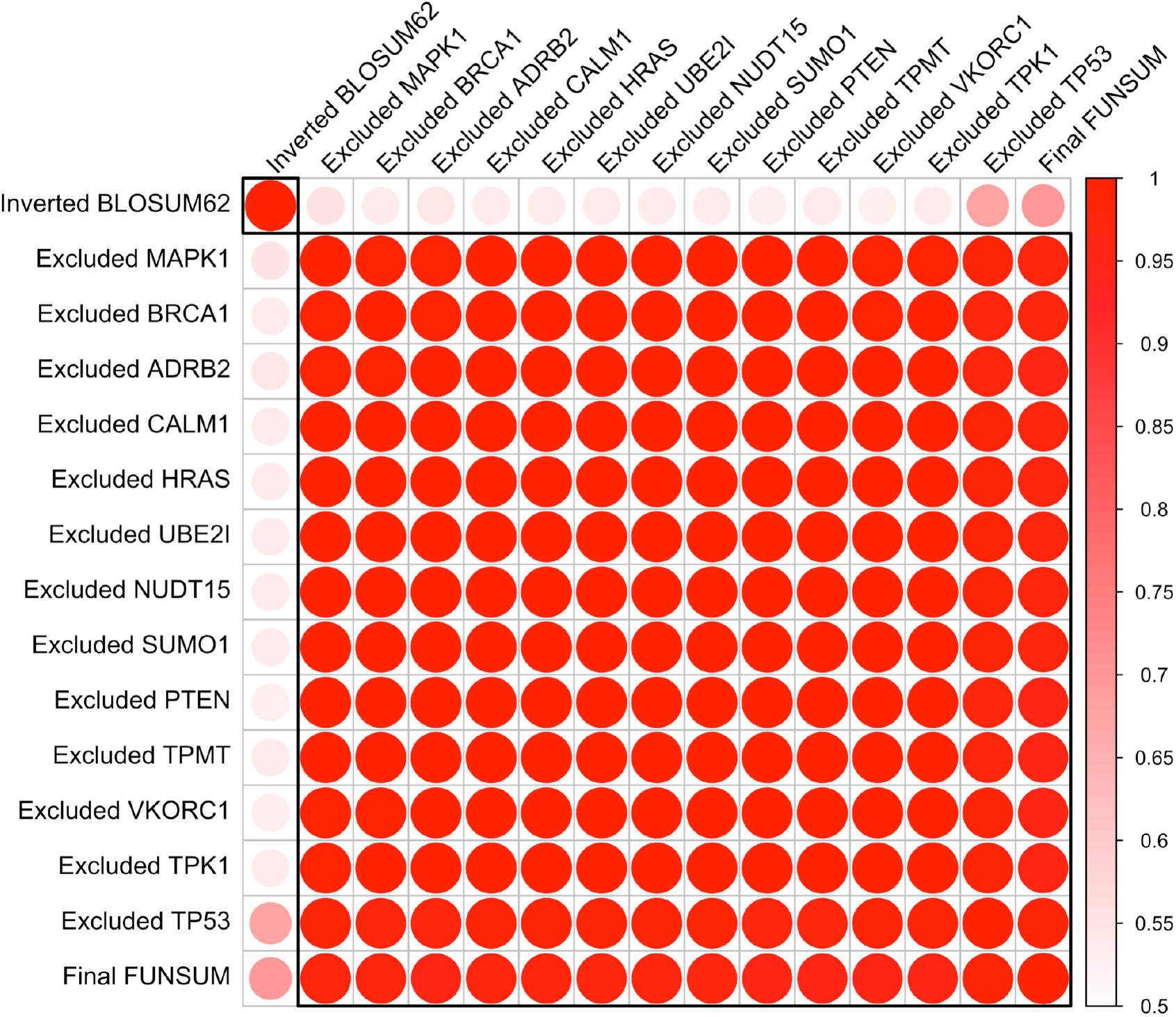
Pearson correlation of various versions of FUNSUM and BLOSUM62. To test the robustness of FUNSUM, alternative FUNSUMs were generated by pooling all available DMS data while excluding data from one gene each time. High correlation was observed among the alternative FUNSUMs and the final FUNSUM, while a lower degree of correlation was observed between the FUNSUM and BLOSUM62 matrices. The BLOSUM62 matrix was inverted to match the direction of FUNSUM.

**Figure S2:**
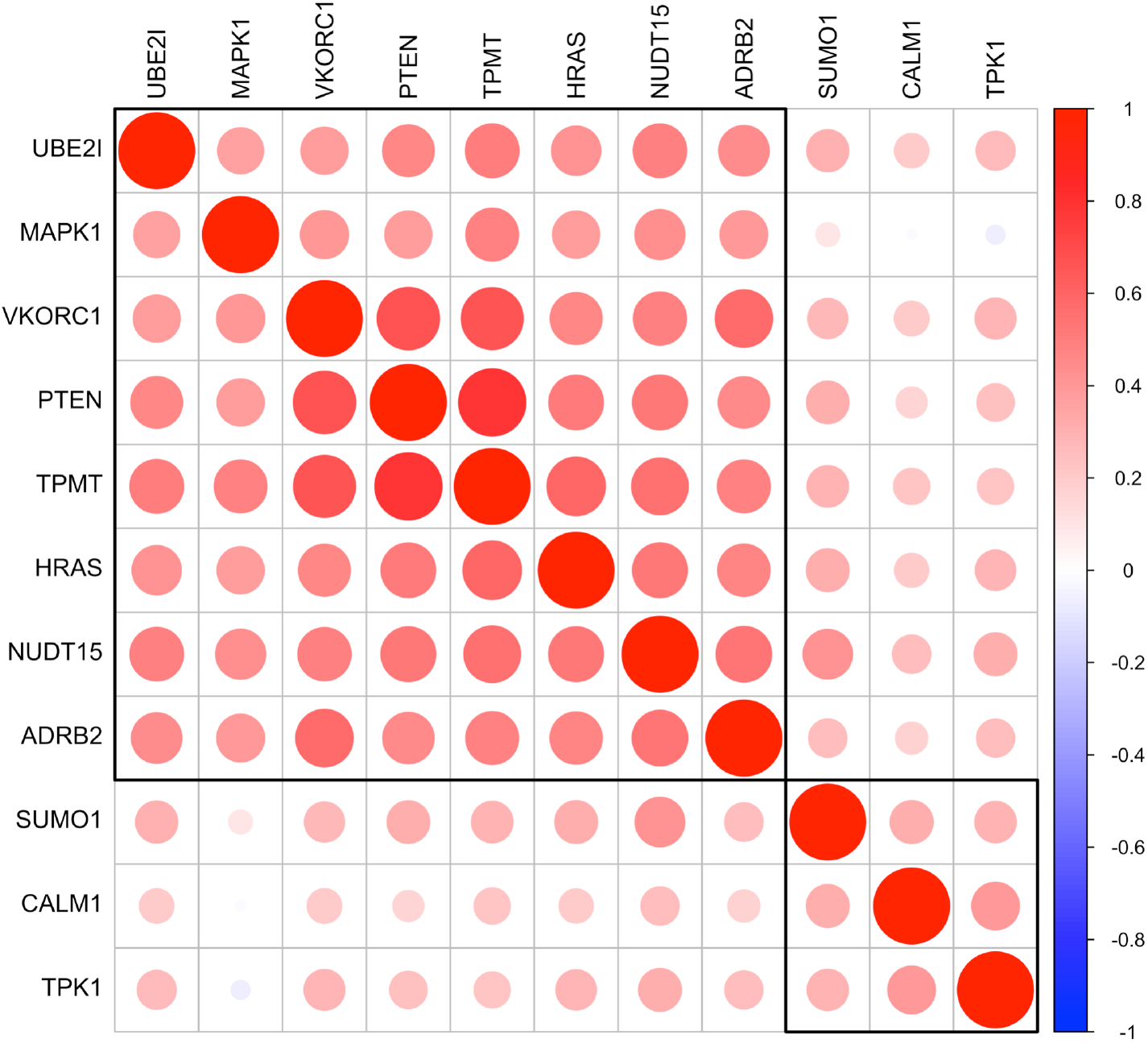
Pearson correlation of FUNSUM generated using DMS data from individual genes. FUNSUM generated from SUMO1, CLAM1 and TPK1 had lower correlation with the FUNSUM generated from the rest of the DMS data, and was therefore excluded from the input DMS data used for generating the final version of the FUNSUM. The black box shows the hierarchical clustering result for 2 clusters.

**Figure S3:**
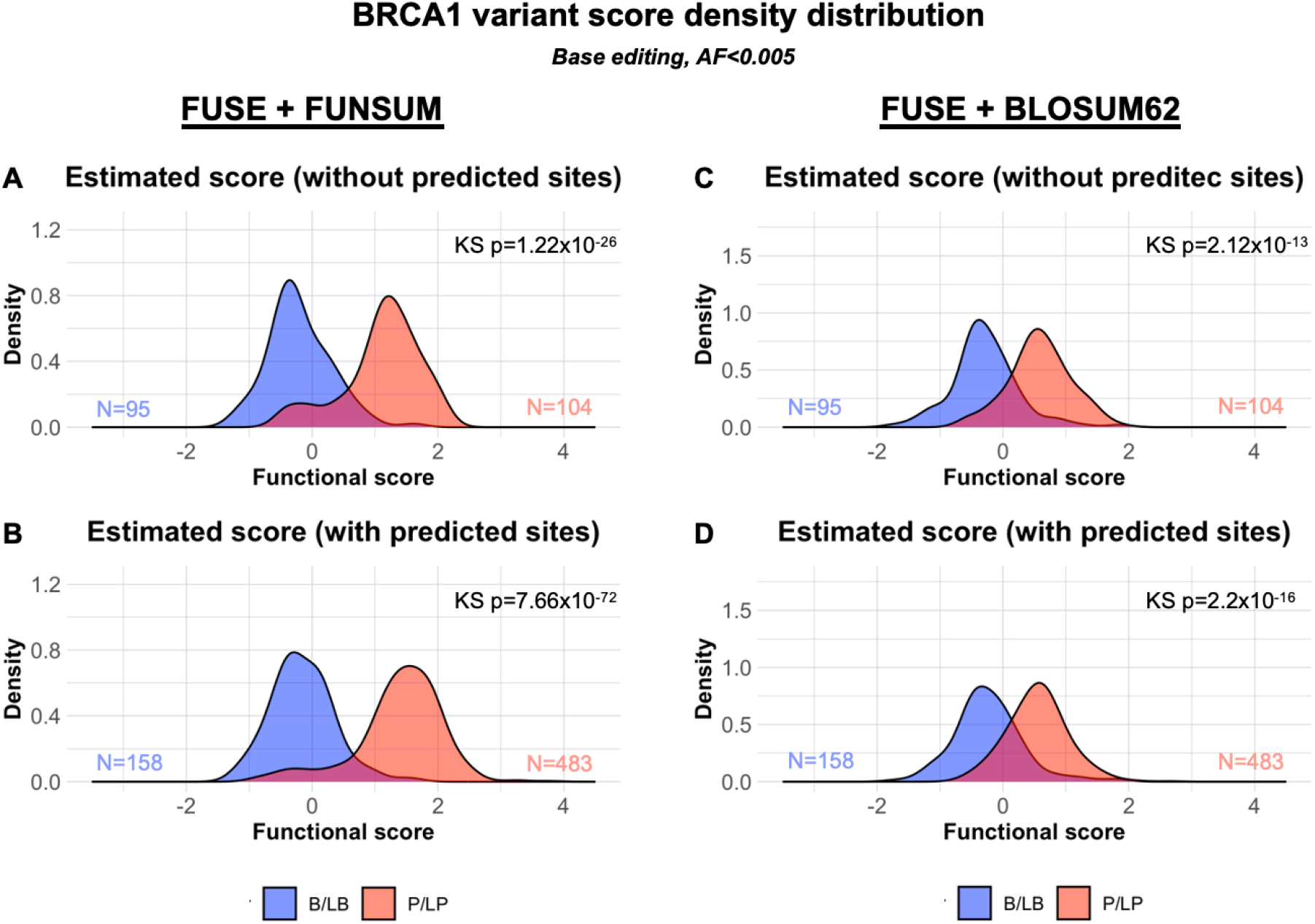
Comparison between FUNSUM and BLOSUM62 in estimating impacts of functional effects using FUSE pipeline. Clinically significant variants were categorized into benign/likely benign group (B/LB, blue) and pathogenic/likely pathogenic group (P/LP, red) based on ClinVar annotations. The numbers of variants included in the B/LB and the P/LP groups are indicated by their corresponding color. For each score and score component, the Kolmogorov–Smirnov (KS-test) was performed to evaluate how much separation can be achieved between the B/LB and the P/LP groups. **[A]** FUSE estimated functional scores for variants in the original assay, using FUNSUM as the substitution matrix. **[B]** FUSE estimated functional scores for variants that were assayed or can be predicted, using FUNSUM as the substitution matrix. **[C]** FUSE estimated functional scores for variants in the original assay, using normalized BLOSUM62 as the substitution matrix. **[D]** FUSE estimated functional scores for variants that were assayed or can be predicted, using normalized BLOSUM62 as the substitution matrix.

**Figure S4:**
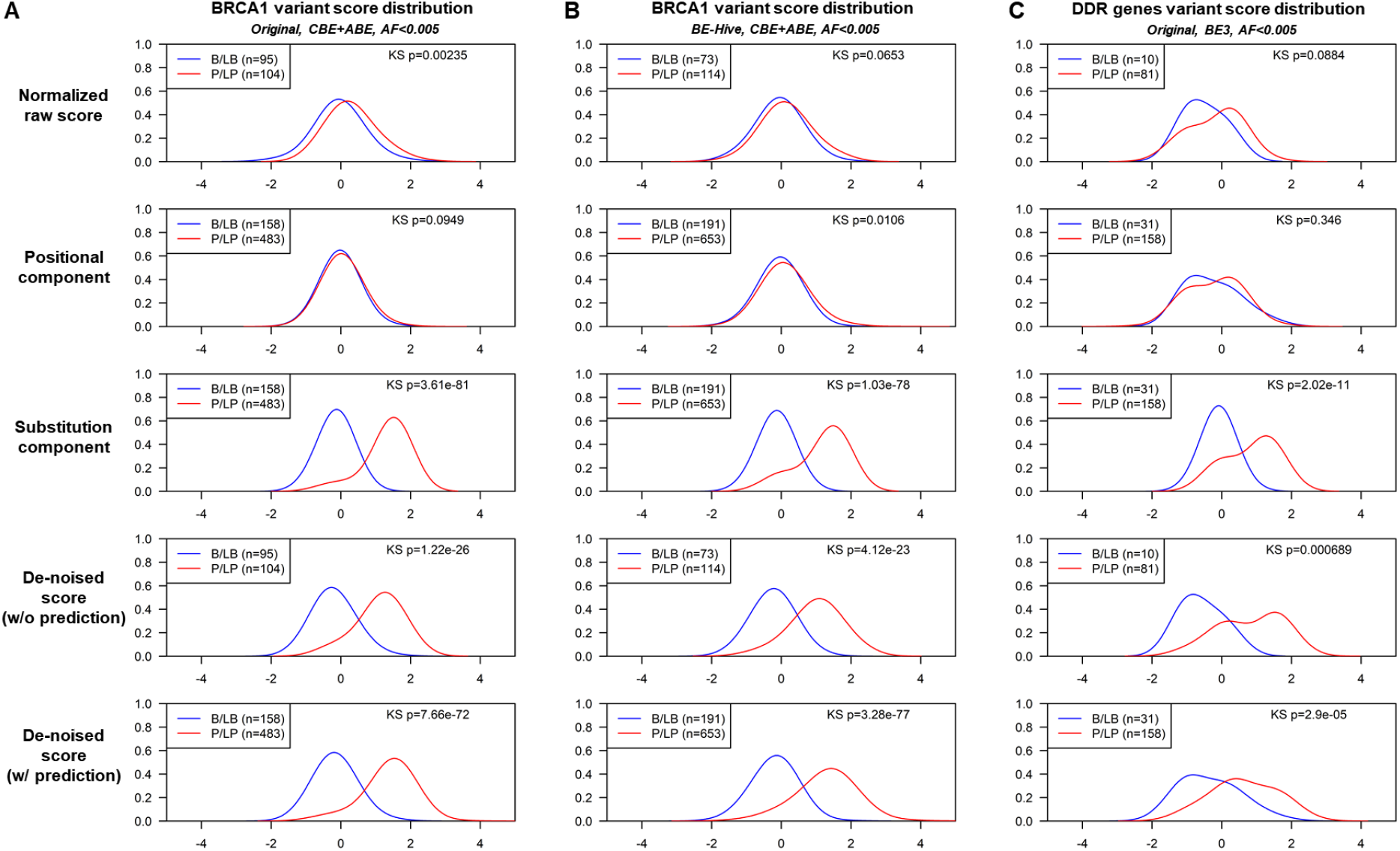
Density plots of predicted functional impact from base editing data for clinically significant variants. Clinically significant variants were categorized into benign/likely benign group (B/LB, blue) and pathogenic/likely pathogenic group (P/LP, red) based on ClinVar annotations. There are three sets of analyses: **[A]** a large base editing assay in BRCA1, **[B]** the same large base editing assay in BRCA1 with substitutions predicted by the BE-Hive algorithm, and **[C]** another large base editing assay in a set of DNA Damage Repair related genes. Within each analysis, there are five row panels: 1) The normalized variant functional scores from the original base editing dataset, 2) the mean positional score as estimated by James-Stein estimation, 3) the predicted impact of the substitution using FUNSUM, 4) the estimated functional score for each variant using the FUSE pipeline, on the originally assayed set of variants, and 5) the estimated functional score for each variant that was assayed or can be inferred using the FUSE pipeline. Numbers of variants in each analysis are included in the legend. For each score and score component, the Kolmogorov–Smirnov (KS-test) was performed to evaluate how much separation can be achieved between the B/LB and the P/LP groups.

**Figure S5:**
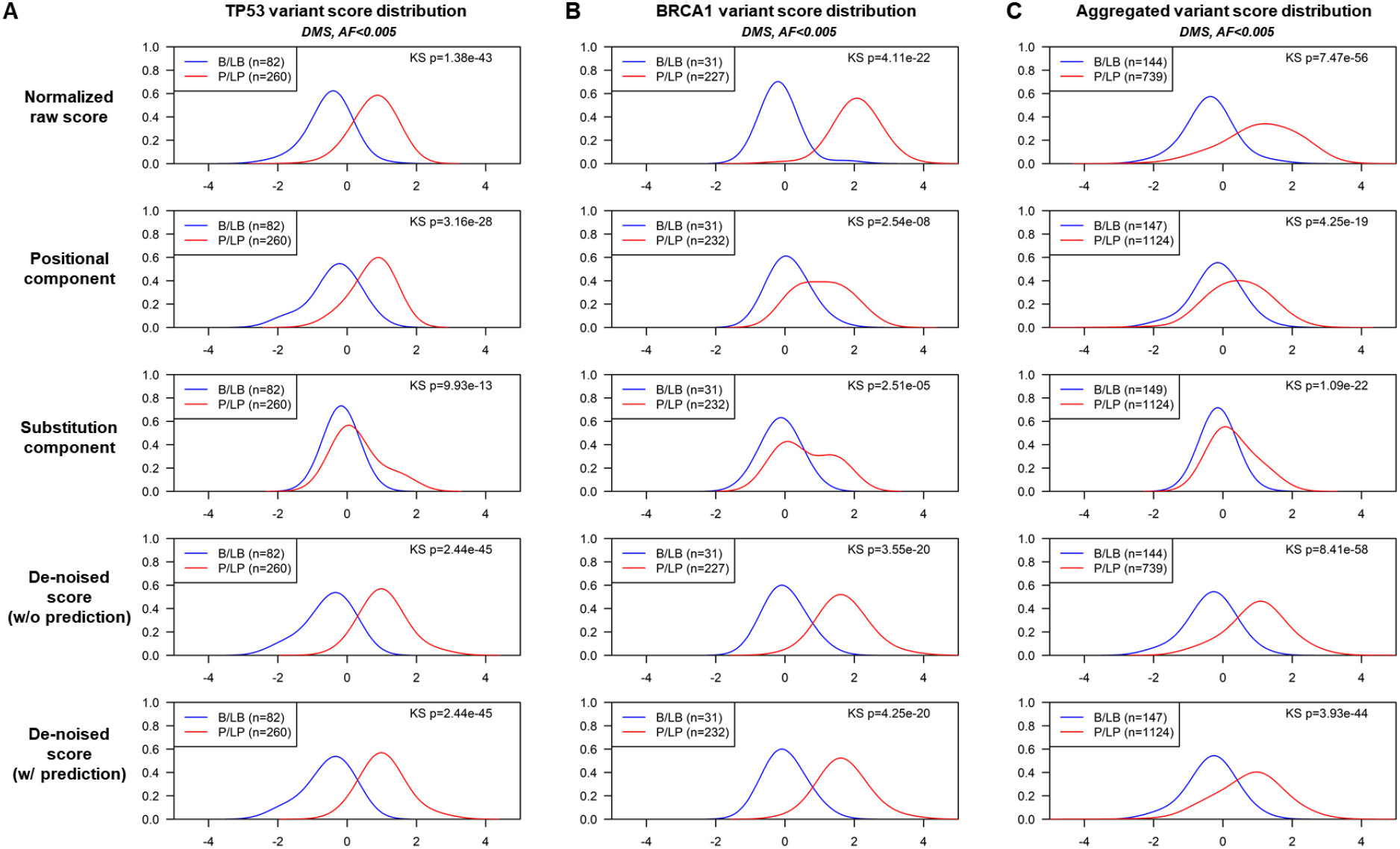
Score density plot for clinically significant variants in deep mutational scanning (DMS) datasets. Clinically significant variants were categorized into benign/likely benign group (B/LB, blue) and pathogenic/likely pathogenic group (P/LP, red) based on ClinVar annotations. There are three sets of analyses: **[A]** a DMS assay in TP53, **[B]** a saturation genome editing assay in BRCA1, and **[C]** a collection of DMS datasets in 10 genes, where each gene’s variant scores were estimated by FUSE with a version of FUNSUM constructed without this gene’s DMS data. Within each analysis, there are five row panels, 1) The normalized variant functional scores from the original base editing dataset, 2) the mean positional score as estimated by James-Stein estimation, 3) the predicted impact of the substitution using FUNSUM, 4) the estimated functional score for each variant using the FUSE pipeline, on the originally assayed set of variants, and 5) the estimated functional score for each variant that was assayed or can be inferred using the FUSE pipeline. Numbers of variants in each analysis are included in the legend. For each score and score component, the Kolmogorov–Smirnov (KS-test) was performed to evaluate the separation between the B/LB and P/LP groups.

**Figure S6:**
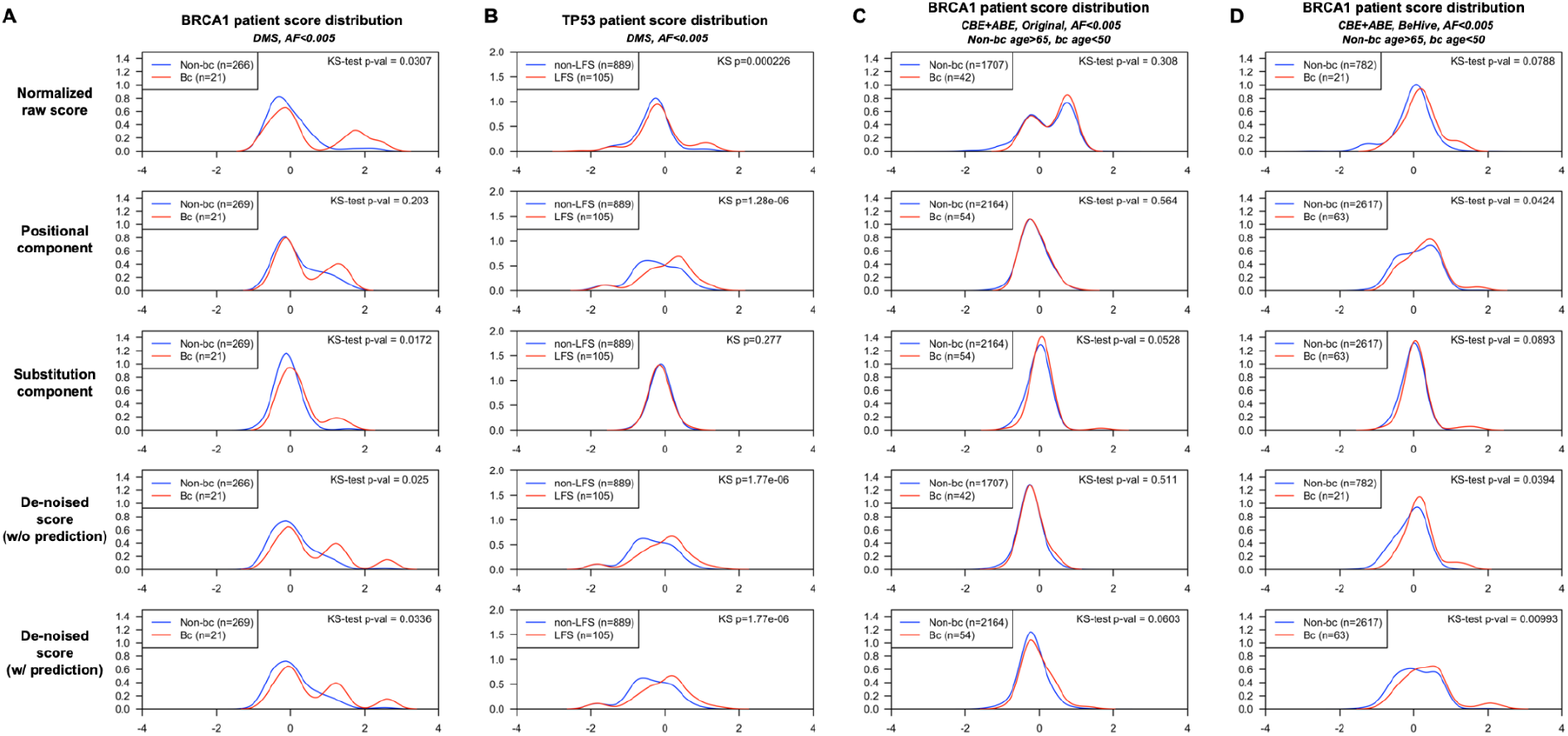
Score density plots for patient groups categorized by UKBB patient characteristics in DMS and base editing datasets. The patient score was calculated as the average variant score of *BRCA1* or *TP53* variants detected in each patient. For *BRCA1*, the patients were categorized by whether they had breast cancer (red), or did not have breast cancer (blue). For *TP53*, the patients were categorized by whether they had Li-Fraumeni syndrome associated cancers (LFS, red), or did not have Li-Fraumeni syndrome associated cancers (Non-LFS, blue). n is the number of patients with a score available. For each score and score component, KS-tests were performed to evaluate the dissimilarity of distributions in the non-cancer group and the cancer groups. **[A]** Patient scores derived from *BRCA1* DMS dataset. **[B]** Patient scores derived from *TP53* DMS dataset. **[C]** Patient scores derived from *BRCA1* base-editing dataset, amino acid changes annotated by the original paper. **[D]** Patient scores derived from *BRCA1* base-editing dataset, amino acid changes predicted by BE-Hive algorithm.

